# Longitudinal testing for respiratory and gastrointestinal shedding of SARS-CoV-2 in day care centres in Hesse, Germany. Results of the SAFE KiDS Study

**DOI:** 10.1101/2020.11.02.20223859

**Authors:** Sebastian Hoehl, Emilie Kreutzer, Barbara Schenk, Sandra Westhaus, Ivo Foppa, Eva Herrmann, India Ettrich, Alexander Schaible, Olga Rudych, Holger Rabenau, Annemarie Berger, Sandra Ciesek

## Abstract

**Background:** With the pandemic of SARS-CoV-2 ongoing in Europe in July of 2020, day care centres were reopened in the state of Hesse, Germany, after the lockdown. The role young children play in the dynamics of the transmission was unknown.

**Methods:** We conducted a longitudinal study over a period of 12 weeks (18^th^ of June 2020 to 10^th^ of September, 2020) to screen attendees and staff from day care centres in the state of Hesse, Germany, for both respiratory and gastrointestinal shedding of SARS-CoV-2. 825 children (age range 3 months to 8 years) and 372 staff members from 50 day care centres, which were chosen representatively from throughout the state, participated in the study. Parents were asked to perform both a buccal mucosa and an anal swab on their children once a week. Staff were asked to self-administer the swabs. RT-PCRs for SARS-CoV-2 were performed in a multiple-swab pooling protocol.

**Findings:** 7,366 buccal mucosa swabs and 5,907 anal swabs were analysed. No respiratory or gastrointestinal shedding of SARS-CoV-2 was detected in any of the children. Shedding of SARS-CoV-2 could be detected in two staff members from distinct day care centres. One was asymptomatic at the time of testing, and one was symptomatic.

**Interpretation:** Respiratory or gastrointestinal shedding of SARS-CoV-2 in children or staff members in day care centres was very rare in the context of low community activity. The data indicate day care centres do not pose a reservoir for SARS-CoV-2 in a low prevalence setting, no inapparent transmissions were observed.

**Funding:** The study was commissioned by the Hessian Ministry of Social Affairs and Integration and was supported by Roche Diagnostics, Basel, Switzerland.

## Introduction

In the pandemic of SARS coronavirus 2 (SARS-CoV-2), pre-school children are at a low risk of severe disease and death when infected with the virus.^1,2^ When social distancing and closure of day care centres were applied to counter the first peak of cases of COVID-19 in Germany in March of 2020, young children were considerably affected by these measures. Restricting access to education is expected to have negative effects on the well-being of children of all age groups, with families of low socioeconomic status and families with children with special educational needs most likely to suffer.^3^ Children are, indeed, believed to be main drivers of the transmission to the community of other respiratory viruses, such as influenza and rhinovirus.^4-6^ But the role pre-school children play in transmission of SARS-CoV-2 was largely unknown when day care facilities reopened after the lockdown, with hygiene restricts in place,^7^ on July 6^th^ of 2020, in the State of Hesse, Germany. Shedding light on the role children play in the dynamics of SARS-CoV-2 transmission ought to be a high priority. Findings from school-age children and adolescents cannot be readily transferred to younger children. Epidemiological data on the unique setting of day care centres are therefore needed.

Since children are often only mildly symptomatic or remain asymptomatic when infected with SARS-CoV-2, it is conceivable that they are shedding virus when attending day care centres, rendering rules that prevent symptomatic children from entering these facilities ineffective. While the viral load of infected children of all age groups was observed not to differ significantly from adults^8^, it has not yet been conclusively demonstrated whether children are as likely as adults to transmit the virus to others or not. Chains of transmission among inapparently infected children could remain undetected without laboratory-based surveillance, and are hypothesized to thusly spill over to the community.^9,10^

To detect SARS-CoV-2 RNA for clinical and diagnosis and surveillance, nasopharyngeal or oropharyngeal swab specimens are commonly recommended. However, both of these sample collection methods need to be obtained by a health care professional using PPE (personal protective equipment), and are very unpleasant for young children, prohibiting their use in longitudinal screenings. Alternative sample collections methods have been proposed to detect respiratory shedding, including saliva samples.^11^ However, supplying these samples may be difficult for young children. Buccal mucosa swab samples can be easily obtained from young children by the guardians, but testing sensitivity and the diagnostic window may both be insufficient to be used as a weekly screening tool without additional tests.^12^ Examining stool or anal swabs samples yields a much longer diagnostic window^13^, but shedding in the stool is not present in all children with respiratory infection.^14,15^ By simultaneously testing for both respiratory and gastrointestinal shedding, however, the diagnostic sensitivity is likely increased.

In the ***SAFE KiDS-Study*** (German: **SA**RS-CoV-2 **F**rüh**E**rkennung in **Ki**tas mit “**D**ual **S**wabs”, English: Early Detection of SARS-CoV-2 in day care centres with “dual swabs”), we enrolled children and staff members from 50 day care centres chosen representatively throughout the State of Hesse, Germany. Participants were invited to be tested weekly for respiratory and gastrointestinal shedding by buccal mucosa and anal swabs for SARS-CoV-2 over a period of 12 weeks to determine whether inapparent chains of transmission could be observed.

Measures to reduce the spread of SARS-CoV-2 in day care centers were put in place by the Hessian Ministry for Social Affairs and Integration and applied during the study period. These included barring children and staff with symptoms of COVID-19, other than runny nose, from entering the facilities, as well as denying access to individuals with known exposure to SARS-CoV-2. Access to the facilities was also denied to children if a household member was symptomatic, or was in quarantine due to contact with SARS-CoV-2. Wearing of masks was not mandatory for children or nor staff. The access of caregivers to the facilities was limited.^7^

## Methods

### Study design

Hesse lies in the centre of Germany, and has 5,993,771 inhabitants, 5·2% (364,226) of which are below the age of 6 years.^16^ A representative sample of day care centres was selected by the State Office of Statistics of Hesse, and the selected facilities were invited to participate in the study. 50 facilities were recruited (figure 1) and 30 participants from each facility, compromising both children and staff members, were invited to provide swab samples once a week. Parents who participated in the study were asked to perform both a buccal mucosa swab as well as an anal swab (“dual swabs”) from their children once a week before visiting the day care centre. They received written instructions and were provided access to a video, available with both English and German subtitles, explaining the goal of the study as well as the swabbing procedure. Written consent was obtained. Providing the swabs was voluntary each week, and parents were instructed not to force sample collection if the child resisted. Participating day care staff were instructed to self-administer the swabs. Samples were collected between 18^th^ of June 2020 to 10^th^ of September 2020.

**Figure 1:**
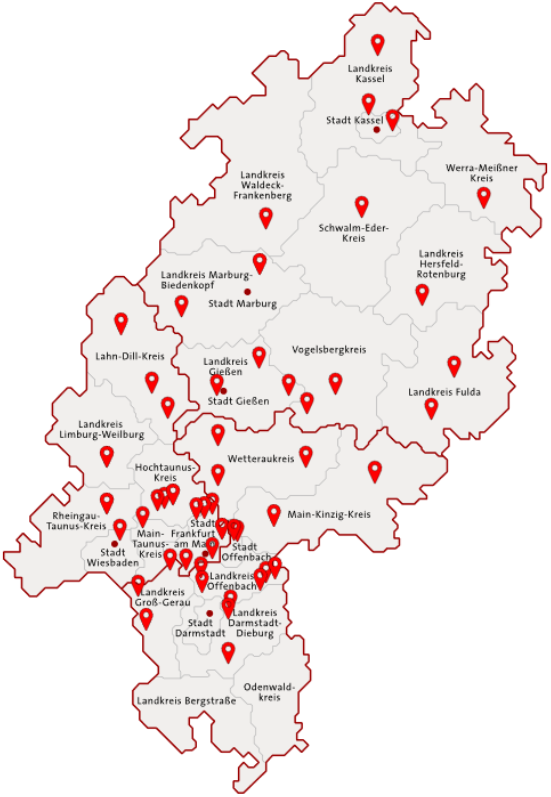
Distribution of participating day care centres throughout in the state of Hesse, Germany. The accumulation of centres in the south of Hesse corresponds to a metropolitan region that has the highest population density of the state.

### Laboratory testing

Testing for SARS-CoV-2 was performed at the Institute of Medical Virology, Goethe University Frankfurt, Germany. Before proceeding to Real-time (RT)-PCR testing, samples were pooled in a 10-sample group-testing mini-pool protocol, that enables efficient use of reagents in a setting with a low pre-test probability, without significant loss in testing sensitivity.^17^ In case of a negative result in the pooled sample, all individual samples receive a negative test result. When at least one of the two PCR targets (E-gene or ORF-region) was detected, all samples of the pool were individually tested. All RT-PCRs were performed on the Roche cobas® 6800 instrument (Roche diagnostics, Basel, Switzerland) according to manufacturer instructions. For all individual samples yielding a positive result for either one or both PCR targets, the public health authority was informed in accordance with the German Infection Protection Act.

To verify the pre-analytic quality of the samples, all of which were obtained without observation by a health-care professional, a sample of 800 buccal mucosa swabs were randomly selected from individual study participants to quantitatively test for GAPDH mRNA by RT-PCR. RNA was extracted with the QIAamp 96 Virus QIAcube HT Kit (QIAGEN, Hilden, Germany) and RT-PCR was performed with the Luna® Universal One-Step RT-qPCR Kit (New England Biolabs, Ipswich, Massachusetts) on the CFX96 Touch Real-Time PCR Detection System (BioRad, Hercules, CA, USA) according to manufacturer’s instructions.

### Questionnaires

At the end of the study, all study participants were asked to fill out a questionnaire to assess of exposure to SARS-CoV-2, and to evaluate whether an infection with SARS-CoV-2 had been diagnosed outside the study. Table 1.

**Table 1:** Numbers of samples that were tested in the study, by age group

| Age of study participant | Number of samples |  |  |
| --- | --- | --- | --- |
|  | Buccal mucosa swab | Anal swabs | total |
| 0 to 2 years | 691 | 634 | 1,325 |
| 3 to 5 years | 3,820 | 3,142 | 6,962 |
| 6 to 8 years | 257 | 214 | 471 |
| No age provided | 173 | 126 | 299 |
| Staff members (age range: 19 to 64 years) | 2,425 | 1,791 | 4,216 |
| **Total** | **7,366** | **5,907** | **13,273** |

### Statistical analysis

The study is analysed in a descriptive statistical assessment because of the low incidence rates. Rates are calculated together with 95% confidence intervals (CI).

### Ethical approval

This study protocol was approved by the ethics board of the University Hospital Frankfurt, Goethe University Frankfurt am Main, Germany.

## Results

### Study participants and sample distribution

A total of 1,197 study participants from 50 day care centres were enrolled in the study. 825 (68·9%) of participants were children, the age range was 3 months to 8 years and 11 months (figure 2). Most children were 4 or 5 years old. 372 participants (31·0%) were day care staff. The age range of staff members was 19 to 64 years. A total of 13,273 valid samples were tested (table 1). 7,366 (55·5%) of these samples were buccal mucosa swabs and 5,907 (44·5%) were anal swabs. 9,057 (68·2%) of samples were from the 825 children, of which 4,941 were buccal mucosa swabs and 4,116 were anal swabs. 4,216 (31·8%) of samples were from the 372 staff members, of which 2,425 were buccal mucosa swabs and 1,781 were anal swabs. The number of samples varied by week (figure 3) and was lower during summer recess, which was from calendar week 28 to 33.

**Figure 2:**
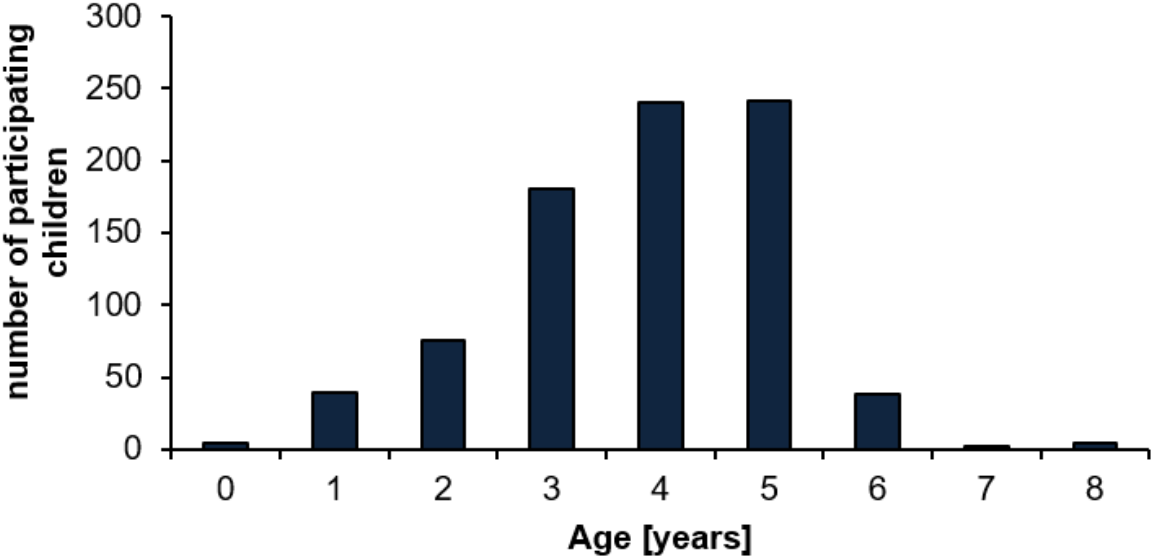
Age distribution of children that participated in the study.

**Figure 3:**
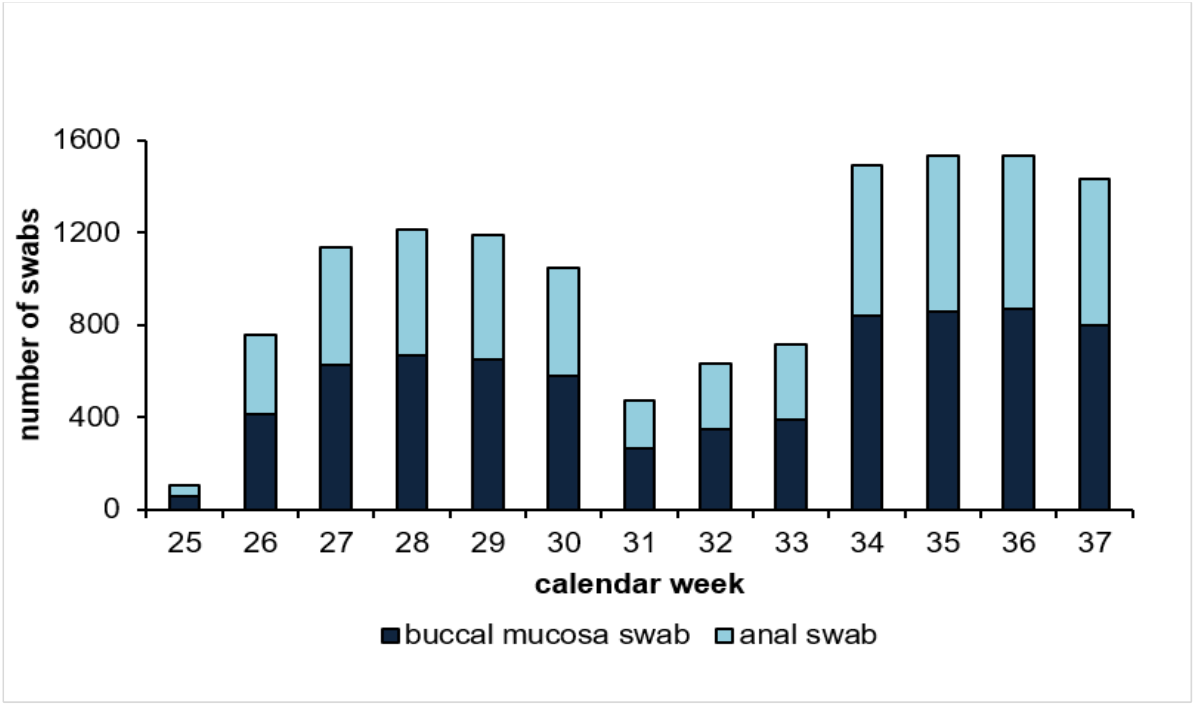
Number of samples provided by study participants, by week. Summer recess occurred from calendar week 28 to 33. Recruitment of centres was not complete in week 25.

### Results of testing for SARS-CoV-2 RT-PCR

Out of 7,366 buccal mucosa swab, SARS-CoV-2 RNA could not be detected in 7,364 (99·97%, 95% CI 99·90% to 100%). Both targets of SARS-CoV-2 RNA (ORF-region and E-gene) were detected in one sample, and only one target gene (E-gene) was detected in one sample. Out of 5,907 anal swab, SARS-CoV-2 RNA was not detected in 5,906 (99·98%, 95% CI 99·90% to 100%).). In one anal swab sample, one out of two targets (E-gene) was detected.

### Description of the detected cases

In calender week 26, SARS-CoV-2 RNA was detected in both the buccal mucosa swab (both PCR targets) and an anal swab (one target) of a day care centre staff member. She was asymptomatic and unaware of the infection at the time of testing. The infection was confirmed by independent testing. The day care center was ordered to be closed for quarantine by the local health department. The local 7 day-incidence of SARS-CoV-2 infections in the administrative distinct of the day care center was low, with 3·5 cases / 100,000 inhabitants, and the 7 day-incidence in children below the age of 8 years was 15·25 cases / 100,000 inhabitants.

In calendar week 34 of 2020, one PCR target (E-gene) was detected in the bucal mucosa swab of a staff member of another day care center. An anal swab was also provided and yielded a negative result. She was sympomatic at the day of testing, and self-isolated after sample collection. A positive result was obtained by independent RT-PCR testing. This day care center was also ordered to be closed for quarantine by the local health department. The total 7 day-incidence of SARS-CoV-2 infections was 35·95 cases / 100,000 inhabitants, and the 7 day-incidence in children below the age of 8 years was 26·18 cases / 100,000 inhabitants.

### Assessment of pre-analytic validity of parent-collected and staff-self-collected buccal mucosa swab samples

800 buccal mucosa swab samples were selected and quantitatively tested for human GAPDH mRNA by RT-PCR, to determine whether material containing cells had been successfully collected. The sample represents 10·9% of all collected buccal mucosa swabs. GAPDH mRNA was detected in 715 (89·4%) of these samples.

### Results from the questionnaires

566 questionnaires were sent in for participating children (return rate 68·6%), and 256 questionnaires were sent in from day care staff members (return rate 68·8%). The questionnaires were anonymously analysed. None of the participants reported to have been diagnosed with a SARS-CoV-2 infection (COVID-19) outside of the study during the study period. Table 2.

**Table 2:** Results of the questionnaire

|  | Children: Caregiver indicating yes (out of 557 participants who returned the questionnaire) | Day care staff: Number of participants indicating „yes“ (out of 256 staff members who returned the questionnaire) |
| --- | --- | --- |
| Was your child / Were you diagnosed with a SARS-CoV-2 infection (COVID-19) during the study period outside of the study? | 0 (0%) | 0 (0%) |
| Was your child / Were you exposed to SARS-CoV-2 (COVID-19) in the household during the study period? | 0 (0%) | 0 (0%) |
| Did your child / Did you have contact with a person with SARS-CoV-2 (COVID-19) during the study period? | 8 (1.4%) | 7 (2.7%) |
| Did your child / did you spend time abroad during the study period? | 118 (20.8%) | 45 (17.6%) |
| *Did your child / did you stay in a country that was designated a “risk area” of COVID-19 by the Robert Koch institute during the study period?* | 5 (0.9%) | 7 (2.7%) |

### Community activity of SARS-CoV-2 in Hesse during the study

During the study period, the incidence of SARS-CoV-2 was relatively low, both for the over-all population as well as for children below the age of 6 years. Regional peaks in the 7-day-incidence in precincst with participating day care centers occurred with up to 66 cases / 100,000 inhabitants.

## Discussion

To our knowledge, this is the first large study examining both respiratory and gastrointestinal shedding of SARS CoV-2 from a representative selection of day care centres during the pandemic.

The overall community incidence of SARS-CoV-2 in the study period was low, but varied by region and study week from 0 to 66 cases / 100,000 inhabitants. In this context, SARS-CoV-2 RNA was detected in none of 7,366 buccal mucosa and 5,907 anal swabs from a total of 825 children attending the day care centres. The absence of detectable respiratory or gastrointestinal shedding of SARS-CoV-2 in any child that participated in our study is reassuring. We could not find evidence for inapparent transmissions of SARS-CoV-2 occurring in the day centres, even though measures recommended to contain transmissions in older children and adults, such as social distancing and wearing of masks, cannot always be applied in the care of young children.

While cases of transmission of SARS-CoV-2 in day care centres to children with subsequent transmission to their household members have been observed^18,19^, the current literature does not indicate that such events are common.^20-23^ As opposed to, e.g., influenza and rhinovirus, where young children and child care facilities likely pose an important reservoir for viral transmissions to the community^6,24^, this does not appear to be the case for SARS-CoV-2, at least in the context of limited community activity.

The only two cases of shedding of SARS-CoV-2 that were detected in our study were in day care staff. Staff members appear to be the likely index case in the clusters that were reported from the day care centres.^18,19^ This is in accordance with a recent study that did not determine an increased risk for day care providers with exposure to child care during the early pandemic in the USA.^25^ While the number of cases detected in staff members in our study was too low to draw conclusions, it should be evaluated further whether screening of staff may be effective in preventing outbreaks of SARS-CoV-2 in day care centres. Our study indicates that screening asymptomatic, young children in a low incidence setting is likely to be ineffective in preventing outbreaks due to the extraordinarily low pre-test probability.

Despite the large number of tests, no single false positive result was observed, demonstrating that the RT-PCR is a highly specific test. Both cases that were detected in our study were later confirmed by independent evaluations. However, we cannot exclude that false negative results may have occurred. The buccal mucosa swab is less sensitive than a nasopharyngeal or throat swab^26^, but could be performed by the caregivers in the study with acceptable pre-analytic quality of the samples. This could be demonstrated by the large amount of buccal mucosa swabs that contained human cells, as determined by the RT-PCR of GAPDH mRNA. In this study, we attempted to increase the sensitivity of the buccal mucosa swab by also examining anal swabs, which yield a longer diagnostic window, especially in young children.^13^ In a survey at the end of the study, none of the participants who returned the questionnaire reported to have received a positive test result outside of the study (table 2), we are therefore not aware of a false negative result having occurred in the study.

This study was conducted in the summer of 2020, when activity of other respiratory pathogens was also low in Hesse, and children with symptoms of upper respiratory infection, other than runny nose only, were excluded from attending day care due to restricts set in place during pandemic. A recent study reported that children without signs or symptoms of COVID-19 rarely tested positive for SARS-CoV-2 RNA, even in a region with a very high burden of COVID-19.^27^ Excluding children with symptoms of respiratory tract infections from attending may be important in reducing the risk of undetected chains of transmission of SARS-CoV-2 in the day care centre setting. This may proof to be a challenge in the coming winter, when upper airway infections in children are expected occur frequently.

A strength of this study was the distribution of participating day care centres throughout Hesse (figure 1), including facilities from both metropolitan and rural regions, with diverse socioeconomic and migration backgrounds, as well as varying activity of SARS-CoV-2. Individual participation in the study by families as well as day care staff was voluntary. This likely caused selection bias, as families who decided to participate in the study may also exhibit a more defensive behaviour in the pandemic. Information on the study and instructions were only provided in German, with an instructional video available with English subtitles, but no other languages were made available. This likely led to an underrepresentation of families with native languages other than German. It is also possible, that the low incidence of SARS-CoV-2 infection reflected the effectiveness of general infection control measures still in effect at the time (mask requirements etc.). Furthermore, in a higher incidence context, a role of young children as transmission vehicles might be more apparent.

In conclusion, we could not detect evidence for inapparent transmission of SARS-CoV-2 occurring in day centres with a local incidence up to 66 cases / 100,000 inhabitants during the pandemic. Further studies should examine whether this is also the case for a setting with higher activity of SARS-CoV-2 infections.

## Data Availability

All relevant data is provided in the manuscript. Additional data will shared upon request.

## Contributions

Sonja Carstens (administrative support); Leon Hildebrand (administrative, technical and graphics support); Franziska Wrobel (administrative and laboratory support); Maria Leondaraki (laboratory support), Avital Reshef (administrative support); Tim Hestermann (administrative support); Felix Schneider (administrative support); Lenny Treeprasertsak (administrative and laboratory support); Wenhan Du (administrative support); Prashidda Khadka (administrative support); Marhild Kortenbusch, Jessica Gille, Regine Jeck (laboratory support); Olga Rudych (administrative support)

## Statistical Consultation

The Hessian State Office for Statistics provided a representative sample of day care centres in Hesse

